# Integrative Approaches for Skin Cancer Detection and Classification : A Dual modal Analysis

**DOI:** 10.1101/2025.08.22.25334277

**Authors:** Nishant Sharma, Manish Kumar Pandey

## Abstract

The early detection of skin cancer is of critical importance, as it can lead to in fatal outcomes if left unaddressed. Given the limited accessibility of dermatological expertise to the general population, it becomes imperative to devise an economical, efficacious, and precise methodology capable of efficient&reliable diagnosis of melanoma and other forms of skin cancer. A data-driven paradigm emerges as the optimal solution for the early and accurate identification of skin malignancies. In this study, we explored an array of Machine Learning (ML) and Deep Learning (DL) algorithms, identifying the most effective model for this context.To conduct this study, we utilized the ISIC 2024 challenge dataset, which contains images of cropped skin lesions, along with metadata associated with each lesion.The developed algorithm is operable without the necessity for specialized clinical intervention and demonstrates exceptional accuracy in differentiating between malignant and benign skin cancer cases, utilizing both cropped skin lesion images from Total Body Photography (TBP) and corresponding tabular data/metadata. For CSV format metadata, Random Forest performed best with an accuracy of 99.928% and an F1-Score of 99.92%, and for cropped skin lesion images, ConViT Tiny performed best with an accuracy of 96.99% and an F1-Score of 96.9%. The output of the proposed work would help in creating a benchmark for preliminary diagnosis of the cancerous tissues and timely prognosis.

## 1 Introduction

Skin cancer is the most commonly diagnosed cancer in the United States. Among its various types, invasive melanoma accounts for only 1% of all skin cancer cases but contributes to the majority of deaths caused by skin cancer [9]. In 2024, an estimated 100,640 new cases of invasive melanoma are projected to be diagnosed in the U.S., with 8,290 individuals expected to succumb to the disease [9].

Early detection of skin cancer is critical, as it significantly improves patient outcomes. The most effective way to detect skin cancer early is by being vigilant about new or changing skin spots. Any new lesions or progressive changes in the appearance of existing ones should be promptly evaluated by a dermatologist or clinician. However, a big population lack access to dermatologic care. In such scenarios, dermoscopy-based Artificial Intelligence(AI) algorithms can play a transformative role by accurately identifying individuals who need clinical evaluation, thus potentially reducing diagnostic delays and significantly impacting early detection. Furthermore, when these dermoscopy-based algorithms are made accessible—such as through mobile applications—they can greatly benefit underserved populations and improve early skin cancer detection, a key factor in enhancing long-term patient outcomes.

Today, the utilization of ML and DL algorithms in the medical domain is one of the most promising solutions for building systems that provide accurate, efficient, and robust tools for the early detection of skin cancer or other cancers where sufficient data is available for training. Skin cancers often start as skin lesions [2]. Among these, melanoma, a type of skin cancer, remains one of the most aggressive and deadly cancers worldwide. Early detection is crucial, as it significantly increases survival rates, yet diagnosing melanoma remains challenging due to the visual complexity and variability of skin lesions. Traditionally, dermatologists rely on clinical examinations and dermoscopy to identify suspicious lesions. However, the effectiveness of these methods is often hindered by factors such as limited access to specialists, human error, and the subjective nature of visual assessments.

One of the key challenges in developing automated systems for melanoma detection is the issue of **class imbalance** in training datasets, where images of benign lesions far outnumber images of malignant lesions. This imbalance can lead to biased models that fail to accurately recognize less common skin conditions. Additionally, the scarcity of large, annotated datasets further complicates the task of training robust and generalizable models.

The goal of this research is to create a benchmark system capable of reliably detecting melanoma, even in resource-limited settings. By developing such automated tools, we envision applications that allow individuals to assess their skin health, enabling earlier detection and timely medical intervention.

## 2 Literature Review

An extensive literature review was conducted to understand the problem from the very basics. The study begins with a systematic review of skin cancer classification using Convolutional Neural Networks(CNNs) in [10], which provides a foundational understanding of how CNNs can be utilized in lesion classification. The paper reviewed various approaches to utilizing CNNs for this purpose. They discussed using pretrained CNNs as feature extractors, followed by a custom classification model like Support Vector Machine(SVM), k-Nearest Neighbor(KNN), or Artificial Neural Network(ANN). They also mentioned using CNNs for end-to-end learning without freezing the classification layer and building the CNN model from scratch, which we implemented in one part of our study. The principal findings of this paper highlighted the absence of a standard benchmark dataset, making it difficult to compare different models. Additionally, the dataset availability and type of data were noted as challenges. For example, the ISIC dataset primarily contains data from individuals with light skin tones, whereas accurate classification requires data from individuals with darker skin tones as well. Overall, this paper provides a general overview of how CNNs can be used in skin lesion classification and the challenges researchers face in building these models.

Another study on the Segmentation and Classification of Skin Cancer Melanoma from Skin Lesion Images in [11] extensively discussed how machine learning algorithms can be utilized in this study. This study uses the dataset provided by ISIC, which we have also utilized in our research. They first performed preprocessing steps such as hair removal from images and noise removal. After this, they used a semi-supervised mean shift segmentation method to segment the lesions from the images. Subsequently, they extracted features to classify the segmented skin lesions.

They extracted features such as those for asymmetry, border, color variation, etc., and then used the RELIEF [13] feature selection algorithm. Feature reduction was performed using linear principal component analysis. After all this preprocessing, segmentation, feature selection, and reduction, they used three ML classifiers: kNN, Decision Tree, and SVM, where SVM achieved the maximum accuracy of 78.2%.

The major critique or challenge of this paper is that they used only 220 training images and 20 test images to train their models and did not provide clear justification for the precision and recall values of the test data. This study could be extended by utilizing deep learning algorithms, which demand more data to yield better results.

One other study on the Detection of Skin Cancer Based on Skin Lesion Images Using Deep Learning [11] emphasizes the utilization of deep learning to accurately and swiftly identify skin cancer. They used the ISIC 2018 dataset and employed CNNs to classify two primary types of tumors: malignant and benign.The authors first used ESRGAN (Enhanced Super-Resolution GAN) to improve the quality of images, which they believe could be beneficial for building a better classification model. They utilized the transfer learning methodology by employing ResNet-50, InceptionV3, and Inception-ResNet.

In the survey paper [15] the authors discussed the challenges faced in skin lesion detection, such as noise and the presence of artifacts, color illumination, and irregular fuzzy boundaries. They also elaborated on preprocessing techniques for skin lesion images, segmentation techniques like conventional intelligencebased methods and deep learning-based techniques, and provided a clear and explainable discussion about the state-of-the-art deep neural networks used in the classification of skin lesion images.The authors discussed CNNs, pretrained CNN models, and convolutional block attention modules. They also highlighted standard skin lesion image databases for model evaluation, such as the ISIC databases, PH2, and HAM10000, which are benchmark datasets in skin lesion image classification tasks. This paper provides a clear perspective on how to proceed with our research study in skin lesion image classification.

In [14], the authors have discussed the methodology of how transfer learning can be utilized to classify cancerous skin lesions from non-cancerous ones. The authors have utilized pre-trained models like VGG-16 & VGG-19 and fused these pre-trained networks, trained on ImageNet, with AlexNet. They then fine-tuned the final architecture with a dermatology image dataset, and the final proposed model achieved an accuracy of 98.18%. The authors have very specifically mentioned the class imbalance problem in medical image analysis and have utilized the ISIC dataset, augmenting it with the Complete-MedNode-Dataset to handle the class imbalance properly.

In [26], the authors have highlighted the potential of AI and DL in classifying various skin conditions. They utilized a dataset with 3,000 images representing nine different skin conditions. They also highlighted the importance of transfer learning in classifying various skin conditions. The authors used VGG16 and ResNet to build a hybrid model, which is more powerful than the individual models, achieving a training accuracy of 98.75% and a validation accuracy of 97.50%. The proposed model has a precision score of 97.60% and a recall score of 97.55%.

A Comparative Study for Classification of Skin Cancer [16] compares the classification results of six different classifiers: SVM (Support Vector Machine), Logistic Regression, Random Forest, AdaBoost, Balanced Bagging, and Balanced Random Forest, in combination with seven feature extraction methods, such as HSV (Hue-Saturation Value), LBP (Local Binary Pattern), HOG (Histogram of Oriented Gradients), and SIFT (Scale Invariant Feature Transform), among others. The study also evaluates four different data preprocessing steps on two skin cancer datasets, ISIC 2016 and HAM10000.The Balanced Random Forest Classifier yielded the best prediction results on the HAM10000 dataset, with an accuracy of 74.75% and an AUC of 81.46%, when the system used Linear Normalization of the input image as the data preprocessing step and HSV as the feature extraction method.

The paper Skin Cancer Classification Using Image Processing and Machine Learning [17] proposed a classification and segmentation technique for skin lesions utilizing image processing and machine learning techniques. They also used the ISIC 2016 dataset.First, they performed image preprocessing, including image resizing, noise removal, and hair removal. Then, they segmented the images using the OTSU thresholding algorithm, followed by feature extraction and reduction. To address the class imbalance in the ISIC dataset, they applied Synthetic Minority Oversampling Technique(SMOTE) [22], resulting in a 50:50 ratio between the minority and majority classes.Finally, they implemented machine learning algorithms such as Quadratic Discriminant, SVM (Medium Gaussian), and Random Forest for classification. Their proposed wrapper method for feature selection, in combination with the Random Forest classifier, achieved promising results compared to other classifiers.

In most of the previous studies, the authors have mainly utilized traditional machine learning approaches, such as models like ANN, SVM, and KNN, while some have used deep learning techniques like CNNs or transfer learning. However, there are very few studies that have utilized recent advancements like the self-attention mechanism or vision transformers, which can provide better classification results than the models used previously. The segregation at the cellular level between malignant and healthy tissues is one aspect, the work in past has not focused on. The proposed work has targeted the differentiation between histologically confirmed malignant and benign cases of skin cancer using cropped skin lesion images from TBP (Total Body Photographs). This differentiation was carried out through the associated metadata in a non-clinical setting, utilizing various machine learning and deep learning algorithms.

## 3 Dataset Description

The study was conducted on the dataset provided by ISIC 2024 Skin Cancer Detection with 3D-TBP challenge on kaggle [8].The images of benign skin lesions (see Fig. 1(a)) i.e. class 0 and malignant skin lesions (see Fig. 1(b))i.e. class 1 are shown in the paper.The Dataset has only two classes so this is a binary classification problem and they have provided two types of data in the challenge:

**Fig. 1:**
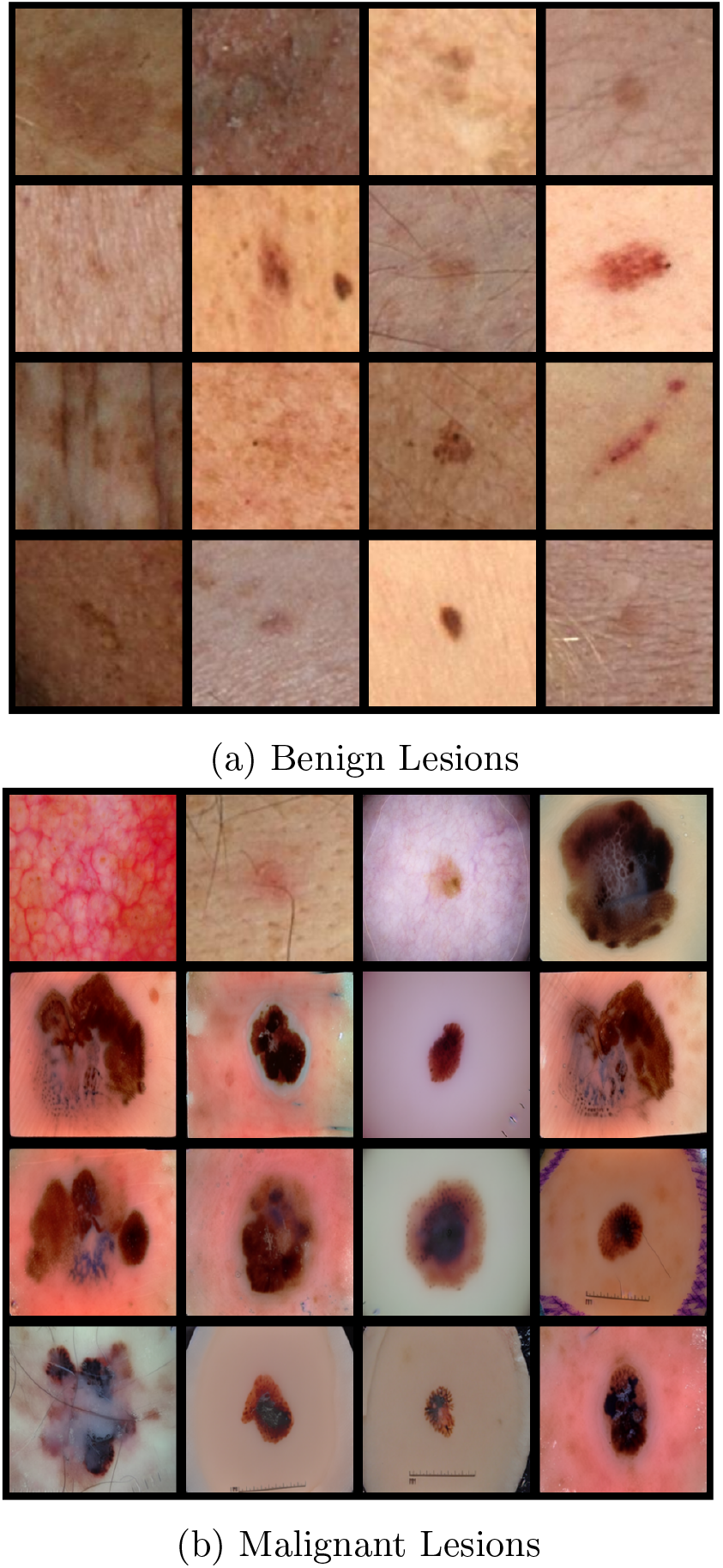
Visual Representation of Benign and Malignant cropped skin lesion images

1. JPEG images of cropped skin lesions
2. Metadata associated with each image in.csv format.

So the study took this challenge as a dual modal problem and experimented with various ML and DL models.The metadata has 55 different features and 393 images that belongs to Malignant class and 401509 images in total.

## 4 Methodology

The experimentation for the proposed work is done in two folds, as represented in the flowchart (see Fig. 2). In the first fold, the image metadata is considered, and in the second fold, the annotated image data is utilized.For the metadata which is in csv format we did experimentation with three ML algorithms i.e. Logistic regression [23], Random Forest [25] and XGBoost [24].We performed the experiments twice once before handling the class imbalance and then after balancing the cases in both classes using SMOTE [22].

**Fig. 2:**
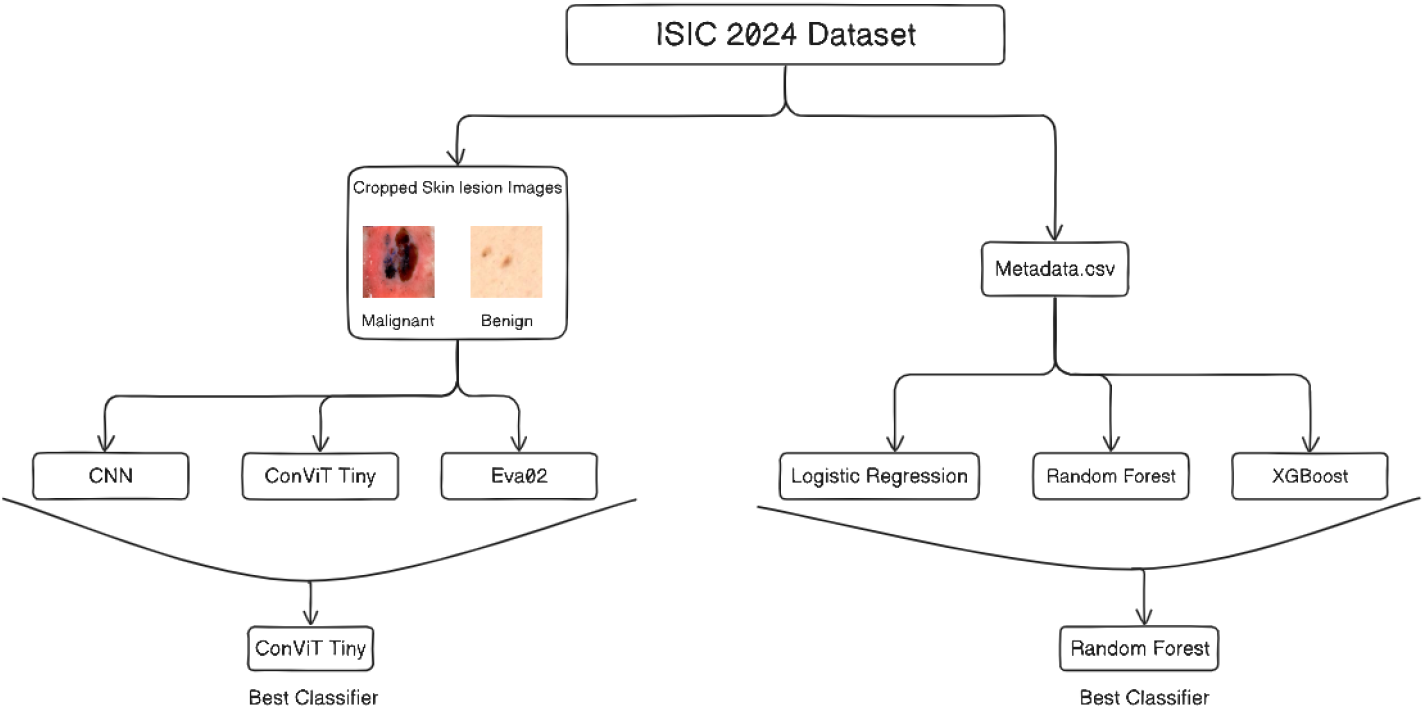
Flow chart for methodology.

We experimented with three start-of-the-art models for the binary image classification task that are listed below :

### 4.1 CNN(Convolutional Neural Network)

CNN’s have been widely used in computer vision tasks particularly in image classification tasks due to their power in capturing local patterns and spatial hierarchies and the translation invariance property. CNNs are comprised of three types of layers. These are convolutional layers, pooling layers and fully-connected layers. When these layers are stacked, a CNN architecture has been formed [18]. This study experimented with different CNN architectures and the proposed architecture gives the best results for lesion classification. Here is the architecture of the model we have used to classify skin lesions(see Fig. 3) :

**Fig. 3:**
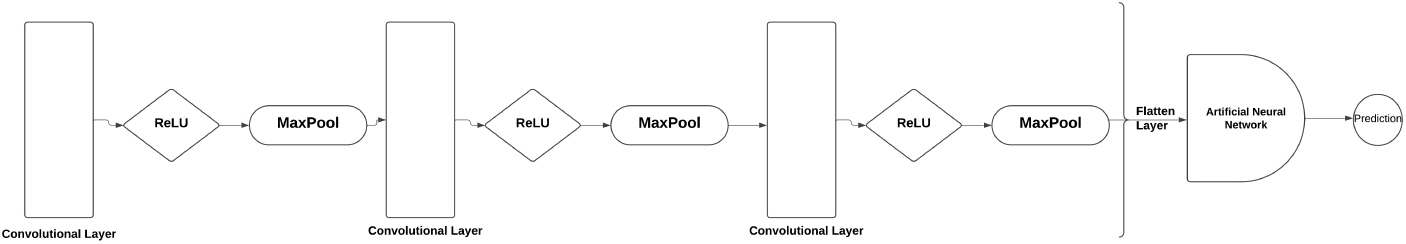
Architecture of the CNN

### 4.2 ConViT(Vision Transformers with Soft Convolutional Inductive Biases)

ConViT [5] is a form of ViT(Vision Transformer) which replaces some of the soft attention layers of ViT with GPSA(Gated Positional Self Attention Layers) which mimic the locality of convolutional layers and give each attention head the freedom to escape locality by adjusting a gating parameter regulating the attention paid to position versus content information [5]. Architecture of the ConViT [5] (see Fig. 4):

**Fig. 4:**
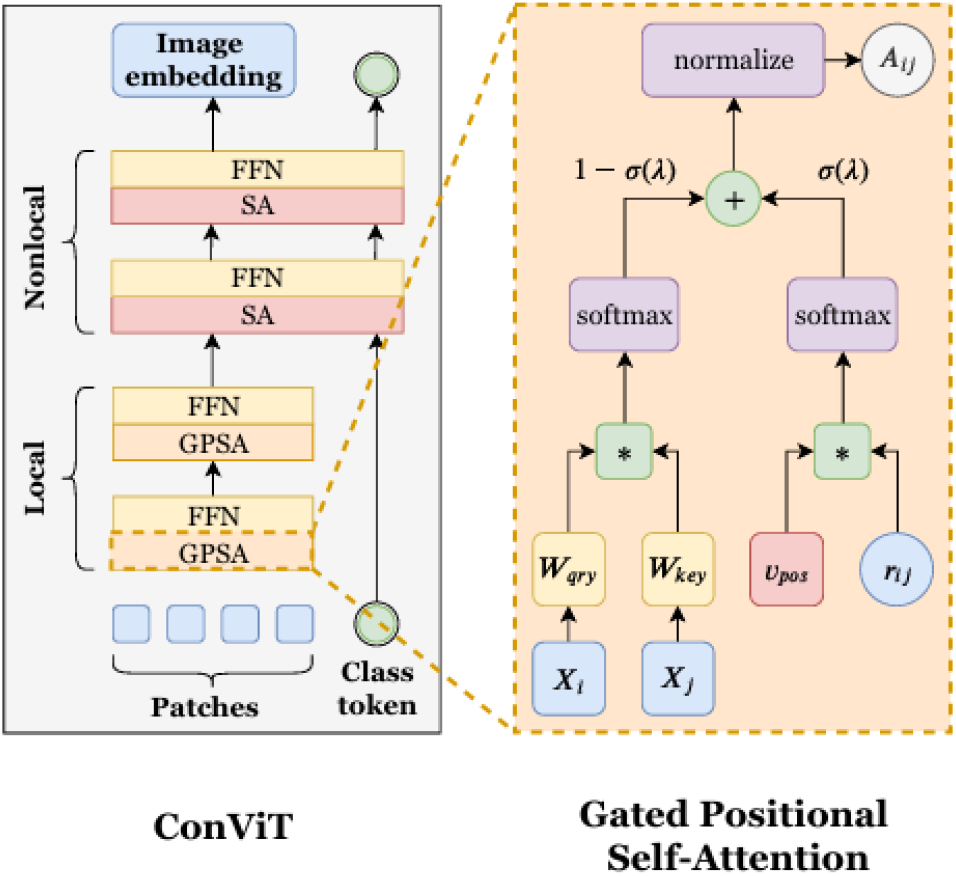
Architecture of the ConViT [5]

### 4.3 Vision Transformer

Vision transformer [20] uses the oiginal encoder of transformer proposed in [21] and then utilizes a classification head to perform classification.The study used EVA-02 [19] (a type of ViT) to perform skin lesion classification because as mentioned in [19] EVA-02 demonstrates superior performance compared to prior state-of-the-art approaches across various representative vision tasks, while utilizing significantly fewer parameters and compute budgets.

## 5 Results and Discussion

At first, the experiments were conducted using metadata in the.csv format. In the Data Pre-processing phase, the null values were checked and handled. Then, categorical variables were one-hot encoded, and the Variance Inflation Factor (VIF) was calculated for each variable. Only variables with VIF<10 were retained. The dataset was then split into training and testing sets with an 80:20 ratio.Finally, standardization was performed (see Equation 1):

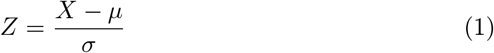

where *µ* is the mean of the training samples and *σ* is the standard deviation.

### 5.1 Evaluation

We used four metrics to evaluate our model’s performance :

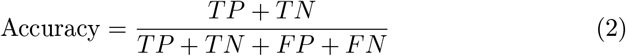

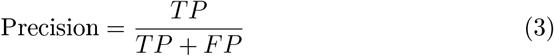

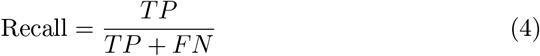

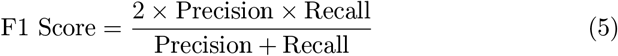

All the experiments were performed twice: once before any class balancing and once after class balancing using SMOTE (Synthetic Minority Oversampling Technique) [22]. There is severe class imbalance in the CSV metadata as well as in the skin lesion image data. This class imbalance was affecting the model’s performance very badly. The majority class has 381533 rows/datapoints, while the minority class has only 381. Consequently, the models are giving absurd results, which cannot be trusted (as seen in the results in Table 2).

**Table 1:** Confusion Matrix.

|  | **Predicted Positive** | **Predicted Negative** |
| --- | --- | --- |
| **Actual Positive** | True Positive (TP) | False Negative (FN) |
| **Actual Negative** | False Positive (FP) | True Negative (TN) |

**Table 2:** Comparison of Model Performance Metrics Before and After SMOTE.

| Model | Before Smote |  |  |  | After Smote |  |  |  |
| --- | --- | --- | --- | --- | --- | --- | --- | --- |
|  | Accuracy | Precision | Recall | F1 Score | Accuracy | Precision | Recall | F1 Score |
| Logistic Regression | 99.882 | 100 | 2.173 | 4.26 | 76.707 | 77.455 | 75.228 | 76.325 |
| Random Forest | 99.879 | 0 | 0 | 0 | 99.928 | 99.914 | 99.942 | 99.928 |
| XGBoost | 99.878 | 0 | 0 | 0 | 99.146 | 99.102 | 99.188 | 99.146 |

To address this issue, SMOTE [22] was performed to balance the cases in the majority and minority classes. We generated synthetic examples of the minority class to make the ratio of majority to minority classes equal using a five nearest neighbors strategy.

After performing SMOTE [22], we reran all three experiments and obtained improved results (see Table 2).

The accuracy of Logistic Regression falls from 99.882 to 76.707, but this model is now better at predicting positive cases (minority classes) than the previous Logistic Regression model, as its F1-score increases from 4.26 to 76.325.

Before performing SMOTE, both Random Forest and XGBoost had zero precision, recall, and F1-score. However, after performing SMOTE, the precision improves from 0 to 99.914, recall improves from 0 to 99.942, and the F1-score improves from 0 to 99.928 for Random Forest. Similarly, for XGBoost, all the metrics (i.e., precision, recall, and F1-score) improve from 0 to 99.102, 99.188, and 99.146, respectively.

After balancing both classes, Random Forest has the best performance metrics among all three models we have used.

In the second fold of experiments annotated images were utilized. Classes were highly imbalanced in ISIC-2024 datset so to handle the class imbalance in images data augmentation was applied.In Data Augmntation the images generated by DERM-T2IM [6] model and the previous ISIC datasets [7] were utilized.

For the images, experiments were conducted with CNN, ConViT, and Eva02 in two phases. In the first phase, experiments were conducted without applying any data augmentation or addressing class imbalance. The results from this phase rendered the model’s ineffective, as the outcomes were highly inconsistent (see Table 3). To address this,additional images were incorporated into the malignant class, which is the minority class, by utilizing data from previous ISIC competitions. This allowed us to balance the ratio of the classes. Subsequently, experiments were ran with all three models across 5, 10, and 50 epochs (see Table 3). At five epochs, ConViT Tiny achieved the highest accuracy and recall, while Eva02 had the highest precision at 98%. ConViT also had a precision of 96%, which is quite commendable. Thus, at five epcchs, ConViT demonstrated strong overall performance (see Fig. 5). At ten epochs, Eva02 achieved the highest accuracy and precision but had a recall lower than ConViT, at 92%. Therefore, at this stage, Eva02 outperformed the other two models (see Fig. 6). At 50 epochs, ConViT exhibited the highest accuracy, precision, and recall (see Fig. 7). However, CNN also performed quite well at this stage, with metrics almost comparable to ConViT.

**Table 3:** Comparison of Model Performance Metrics Before and After Class Imbalance.

|  |  | Before Data Augmentation |  |  |  | After Data Augmentation & Class Balance |  |  |  |
| --- | --- | --- | --- | --- | --- | --- | --- | --- | --- |
| Model | Epochs | Accuracy | Precision | Recall | F1 Score | Accuracy | Precision | Recall | F1 Score |
| CNN | 5 | 50.03 | 0 | 0 | 0 | 90.59 | 96.372 | 84.36 | 89.96 |
| CNN | 10 | 99.62 | 0 | 0 | 0 | 91.56 | 94.503 | 88.244 | 91.26 |
| CNN | 50 | 99.93 | 0 | 0 | 0 | 96.52 | 97.70 | 95.285 | 96.47 |
| ConViT-Tiny | 5 | 99.96 | 0 | 0 | 0 | 95.36 | 96 | 95 | 95.497 |
| ConViT-Tiny | 10 | 99.97 | 0 | 0 | 0 | 94.86 | 92 | 98 | 94.90 |
| ConViT-Tiny | 50 | 99.95 | 0 | 0 | 0 | 96.99 | 98 | 96 | 96.98 |
| Eva02 | 5 | 99.94 | 0 | 0 | 0 | 94.04 | 98 | 89 | 93.28 |
| Eva02 | 10 | 99.95 | 0 | 0 | 0 | 94.23 | 96 | 92 | 93.95 |
| Eva02 | 50 | 99.93 | 0 | 0 | 0 | 95.24 | 97 | 93 | 94.95 |

**Fig. 5:**
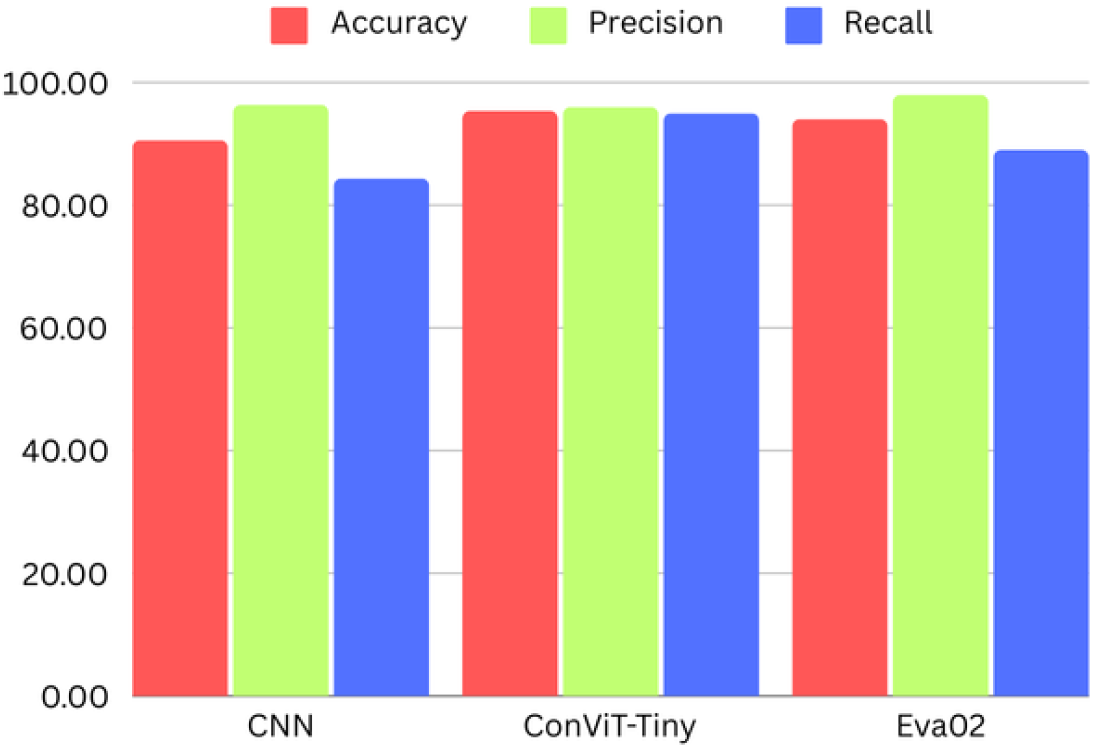
Comparison of model’s performance metrics at five epochs

**Fig. 6:**
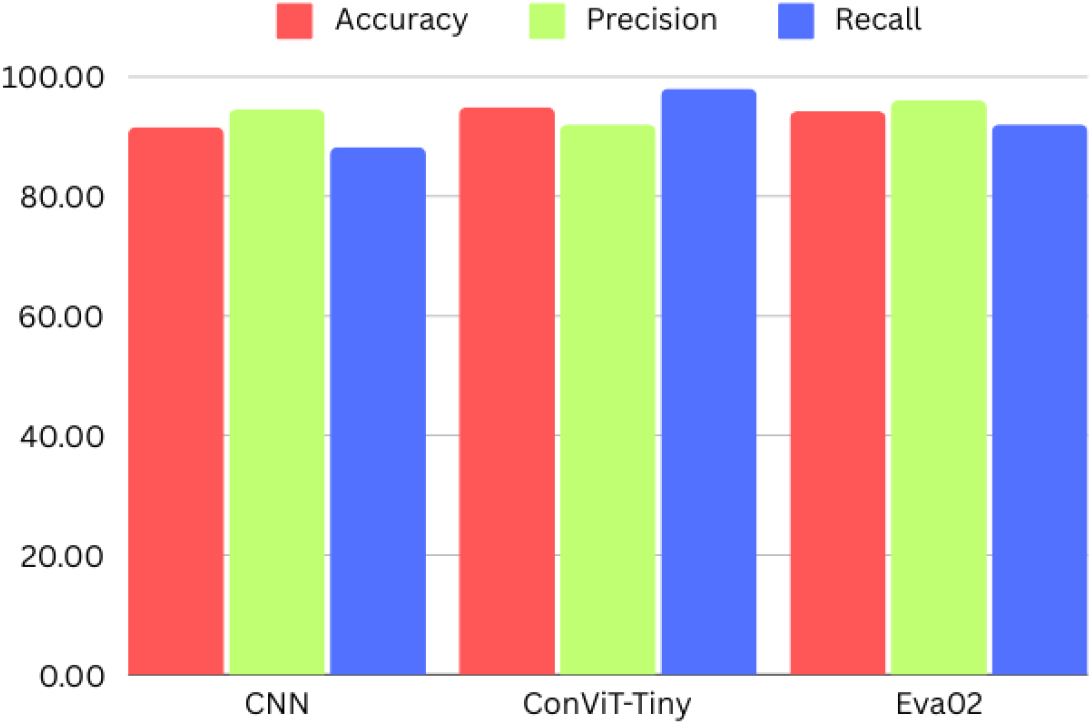
Comparison of model’s performance metrics at ten epochs

**Fig. 7:**
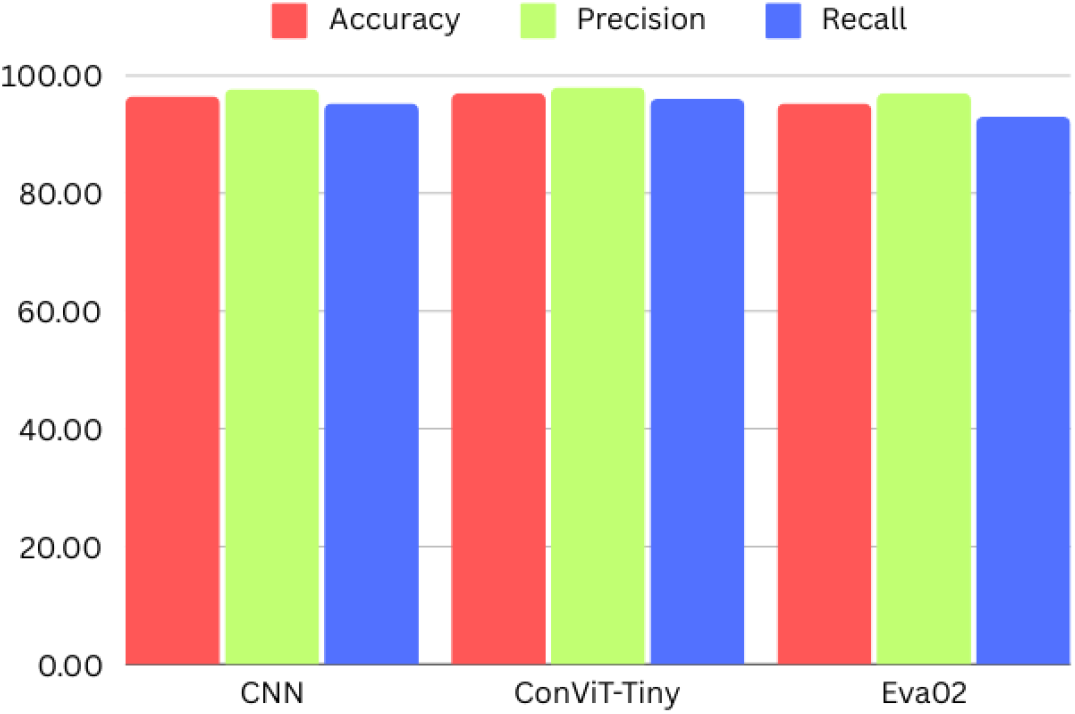
Comparison of model’s performance metrics at fifty epochs

Figures 8, 9, 10 illustrates how the performance metrics and loss vary as the number of epochs increases to 50.

**Fig. 8:**
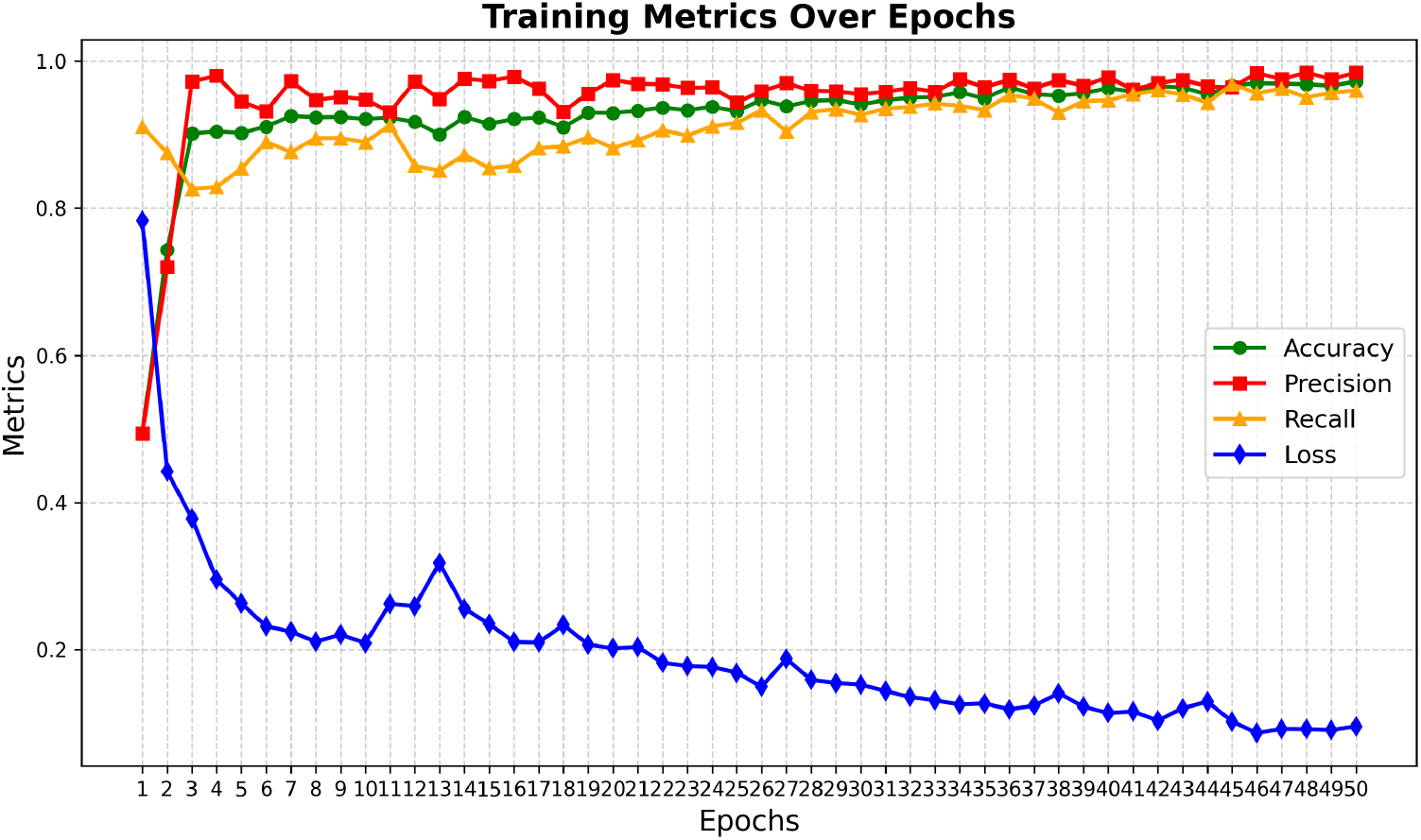
CNN 50 Epochs

**Fig. 9:**
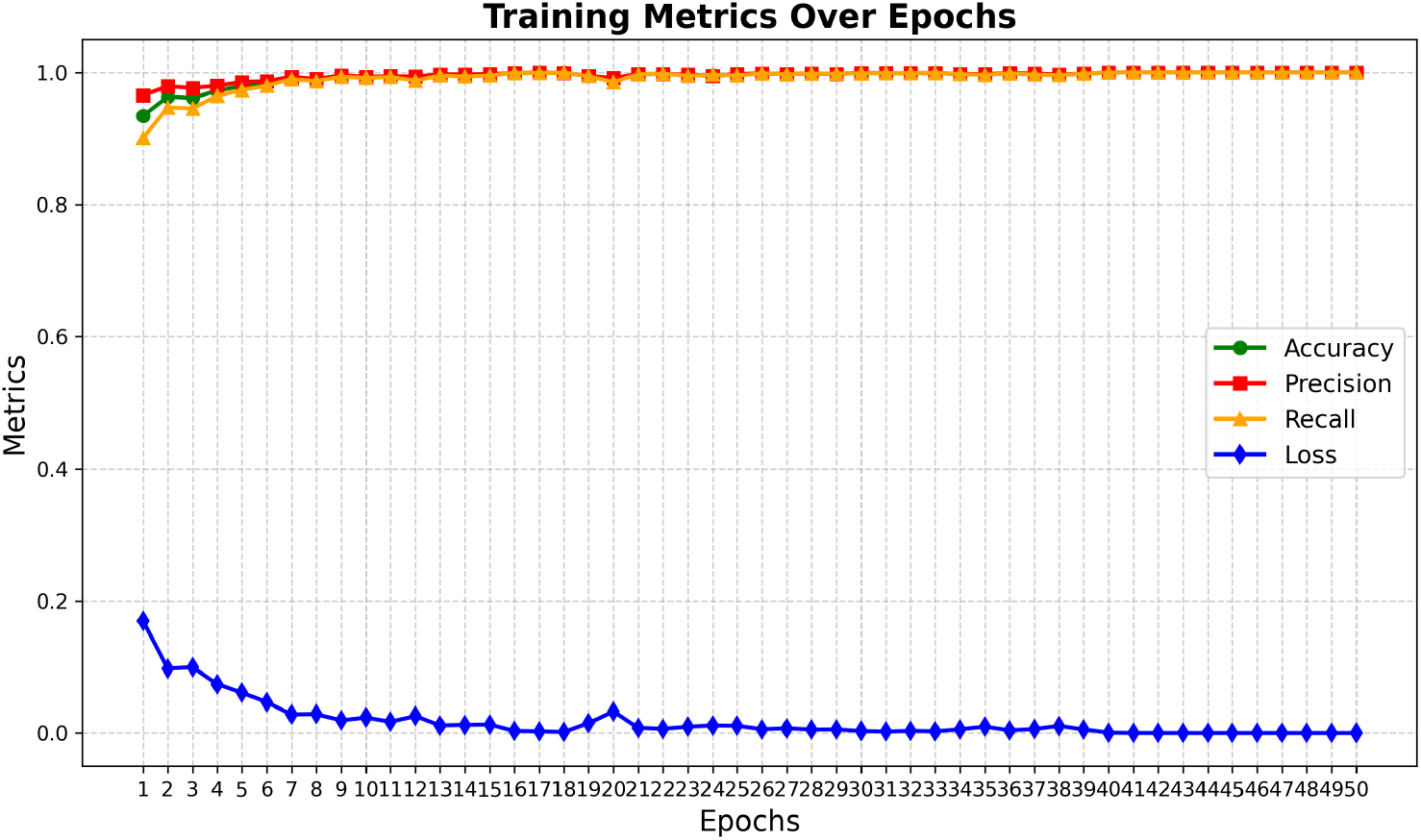
ConViT 50 Epochs

**Fig. 10:**
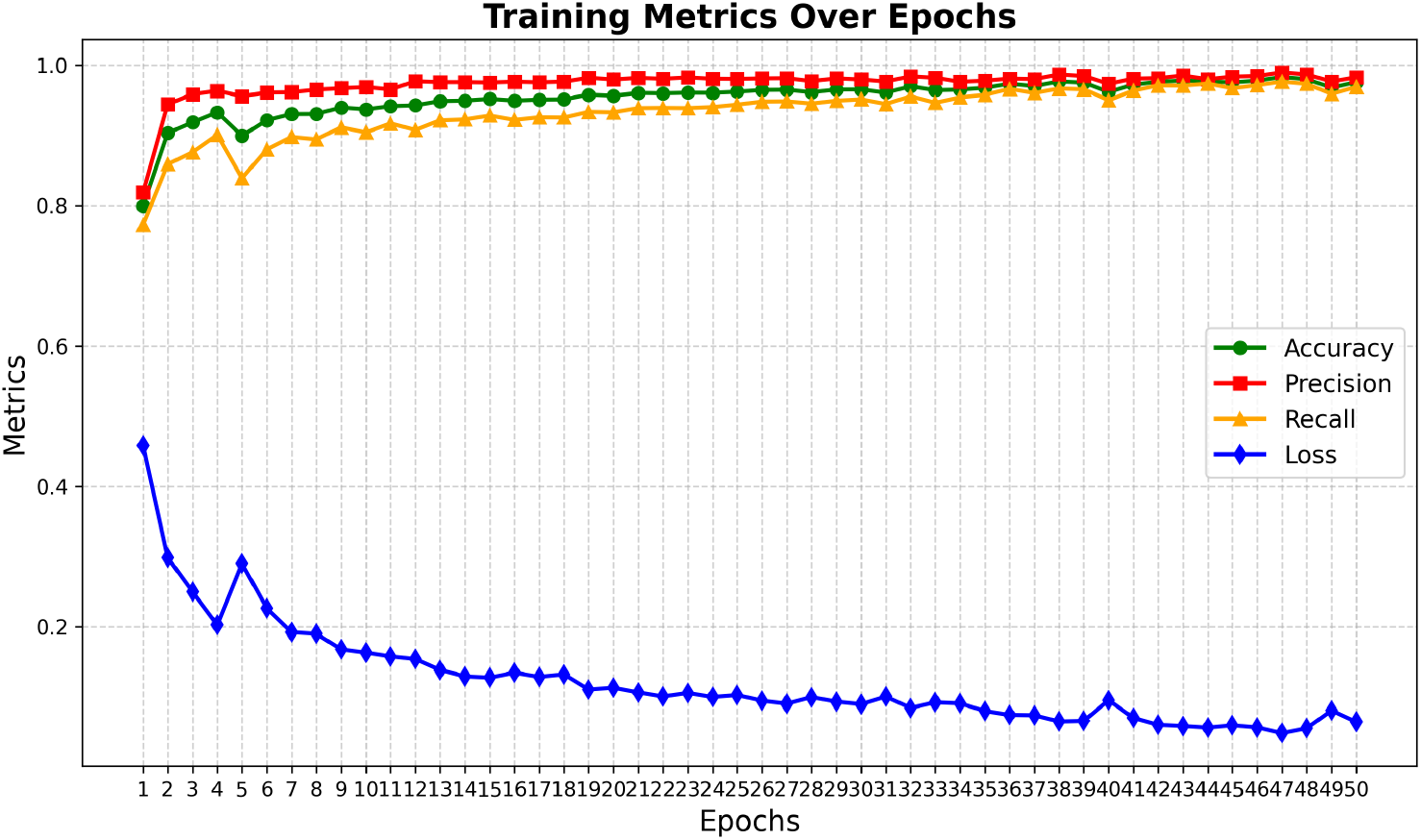
EVA_02 50 Epochs

## 6 Conclusion & Future Work

After conducting these extensive experiments, One can conclude that for skin lesion image classification, ConViT performs better than the other two models. The replacement of dot product self-attention with GPSA (Gated Positional Self-Attention) [5] in certain Transformer layers proves to be highly effective in real-world scenarios, such as skin lesion classification. However, as the number of epochs increases from 5 to 10 to 50, traditional Convolutional Neural Network (CNN) models also show competitive performance, achieving accuracy levels close to those of ConViT and for the metadata associated with each image, the Random Forest model performs the best among Logistic Regression, Random Forest, and XGBoost. The major challenge faced during this study was class imbalance and the insufficient number of images to adequately train the models. In the future, diffusion models can we utilized to generate images for the minority class, helping to balance the datasets and improve model generalization.This study, utilizes skin lesion images generated by the Derm-T2IM model [6], but did not develop its own model. Therefore, future work could focus on experimenting with the development of our own generative model to create more accurate, efficient, and robust models for melanoma detection.

One real-world application could involve developing a mobile application that utilizes these models, enabling individuals to assess their risk of melanoma and take preventive measures in advance. This could be especially impactful for people in underserved or remote areas where access to dermatologists is limited. However, this would require further collaboration with healthcare professionals to ensure that the app’s predictions are both reliable and clinically actionable.

While progress is there, there are still limitations, such as the reliance on a pre-trained model and challenges in obtaining a sufficiently diverse dataset. To address these, we can focus on gathering more data from different sources and exploring techniques like few-shot learning or exploring other generative models (e.g., GANs or VAEs) to generate high-quality synthetic images for the minority classes. Another area that needs to be explored is Explainable AI (XAI) i.e. model interpretability and reasoning in early skin cancer prediction, where the model should be able to explain why a particular image or case has been predicted as positive or negative.

## Data Availability

Dataset is openly available at: ISIC 2024 challenge.

https://challenge2024.isic-archive.com/

